# Seroprevalence and molecular characterization of viral hepatitis and HIV co-infection in the Central African Republic: Viral hepatitis and HIV co-infection in the Central African Republic

**DOI:** 10.1101/2023.08.24.23294539

**Authors:** Parvine Basimane-Bisimwa, Giscard Wilfried Koyaweda, Edgarthe Ngaïganam, Ulrich Vickos, Ornella Anne Demi Sibiro, Brice Martial Yambiyo, Benjamin Seydou Sombié, Pulchérie Pélembi, Sandrine Moussa, Claudine Bekondi, Tamara Giles-Vernick, Alexandre Manirakiza, Muriel Vray, Narcisse Patrice Joseph Komas

**Affiliations:** Viral hepatitis laboratory, Institut Pasteur de Bangui, PO Box 923, Bangui, Central African Republic; Université Evangélique en Afrique (UEA), Bukavu, Democratic Republic of Congo; International Center Advanced for Research and Training (ICART), Bukavu, Democratic Republic of Congo; Hôpital Général de Référence de Panzi, Bukavu, Democratic Republic of Congo; Laboratory of arbovirus, haemorrhagic fevers, emerging virus and zoonosis, Institut Pasteur de Bangui, PO Box 923, Bangui, Central African Republic; Epidemiology service, Institut Pasteur de Bangui, PO Box 923, Bangui, Central African Republic; Centre National de Recherche et de Formation sur le Paludisme, Ouagadougou, Burkina Faso; Service des Retrovirus-VIH, Institut Pasteur de Bangui, PO Box 923, Bangui, Central African Republic; Centre de Ressources Biologiques, Institut Pasteur de Bangui, PO Box 923, Bangui, Central African Republic; Anthropology & Ecology of Disease Emergence Unit, Institut Pasteur, Paris; Unit of epidemiology of emergent infections, Institut Pasteur, INSERM, Paris

**Keywords:** HBV, HCV, HBV-HDV co-infection, HIV-HBV coinfections, Prevalence, HBV/HDV Molecular characterization, Hepatitis B virus, Hepatitis C virus, Hepatitis D virus, CAR

## Abstract

**Background:** The Central African Republic (CAR) is one of the countries with the highest prevalence of viral hepatitis infection in the world. The present study describes the geographic distribution of viral hepatitis infections and molecular characterization of these viruses in the CAR.

**Methodology:** Out of 12,599 persons enrolled during the 4th Multiple Indicator Cluster Survey of 2010 in the CAR, 10,621 Dried Blood Spot (DBS) samples were obtained and stored at -20°C. Of these DBS, 4,317 samples were randomly selected to represent all regions of the CAR. Serological tests for hepatitis B, D, and C viruses were performed using the ELISA technique. Molecular characterization was performed to identify strains.

**Results:** Of the 4,317 samples included, 53.2% were males and 46.8% females. The HBsAg prevalence among participants was 12.9% and that HBc-Ab was 19.7%. The overall prevalence of the HCV was 0.6%. Co-infection of HIV/HBV was 1.41% and that of HBV/HDV was 16.6%. A total of 77 HBV, 6 HIV, and 6 HDV isolates were successfully sequenced, with 72 HBV (93.5%) strains belonging to genotype E and 5 (6.5%) isolates belonging to genotype D. The 6 HDV isolates all belonged to clade 1, while 4 recombinants sub-type were identified among the 6 isolates of HIV.

**Conclusion:** Our study found a high prevalence of HBV, HBV/HDV and HBV/HIV co-infection, but a low prevalence of HCV. CAR is still in an area of high HBV endemicity. This study’s data and analyses would be useful for establishing an integrated viral hepatitis and HIV surveillance program in the CAR.

## Introduction

Hepatitis is defined as an inflammation of the liver. It can be self-limiting or in some cases, progress to fibrosis (scarring), cirrhosis, or liver cancer. Hepatitis may be caused by toxins, certain drugs, heavy alcohol use, and bacterial and viral infections. Hepatitis viruses are the most common cause of hepatitis in the world [1].

There are 5 main hepatitis viruses, referred to as types A, B, C, D, and E. Each type of hepatitis is caused by a different virus, is spread differently, and accounts for variable prevalence and mortality around the world [1]. Globally, viral hepatitis infection is a major public health problem.Each year, 1.4 million people die from viral hepatitis-related cirrhosis and liver cancer [2]. Prevalence varies according to geographical area. Following Asia, Africa has the second largest number of chronic HBV carriers and is considered to be a region of high endemicity [3]. In 2015, the number of deaths caused by hepatitis viruses resembled the number of deaths caused by tuberculosis and higher than those caused by HIV [4]. Viral hepatitis mortality has been increasing over time, whereas tuberculosis and HIV mortality has declined. In most cases, mortality due to viral hepatitis in 2015 was caused by chronic liver disease and primary liver cancer, causing some 720,000 and 470,000 deaths respectively [4]. The HCV epidemic affects all regions of the world, with major differences between and within countries [4]. The HDV, a satellite RNA-virus depending on HBV for its propagation. Both HBV and HDV occur worldwide, with an estimated 240 million people worldwide chronically infected with HBV [4,5], of whom approximately 15 to 20 million, or 5% are co-infected with HDV [6]. In combination with HBV, HDV causes the most severe form of viral hepatitis in humans, including fulminant hepatitis and hepatocellular failure, with rapid progression to hepatic cirrhosis followed by hepatic decompensation, and an increased risk of hepatocellular carcinoma [7–9]. Additionally, HBV/HIV coinfection increases the morbidity and mortality beyond the cause of mono-infection by either one of those viruses. HBV patients coinfected with HIV have higher levels of hepatitis B viremia, have progression to chronic hepatitis B that is approximately five times as fast as that among people infected with only HBV, and have a higher risk of cirrhosis and hepatocellular carcinoma[10].

Although Africa has very high HBV endemicity, HDV prevalence remains poorly known and is thus increasingly studied [4]. Nevertheless, in Africa, of the estimated 65 million chronic HBV carriers, about one-fourth of HBsAg-positive individuals show dual-infection with HDV. Thus, Central Africa has an overall prevalence of 25.6%, West Africa 7.3%, and East and South Africa 0.05% [11].

Numerous seroprevalence studies of viral hepatitis have been conducted in CAR between 1984 and the present day, most focusing on the capital Bangui and its suburbs [12,13]. Although national studies to evaluate national prevalence of HIV (3.5%) [14] and HIV/HBV coinfection have been conducted, in addition to Bangui-centered studies of HBV/HDV [11,15], no studies to date have estimated CAR’s HBV/HDV national prevalence. This study therefore sought describe the national geographic distribution of viral hepatitis infections and of HBV/HIV and HBV/HDV co-infections, and to produce a molecular characterization of these viruses among CAR’s general population.

## Methodology

### Study strategy and sample size

This is a non-interventional descriptive (retrospective) study based on Dried Blood Spot (DBS) samples obtained as part of the 4th Multiple Indicator Cluster Survey (MICS4) in the Central African Republic (CAR) in 2010, carried out as part of the United Nations Children’s Fund (UNICEF) program to estimate the prevalence of HIV infection in the country’s general population. Out of the 12,599 persons enrolled during this MICS4, 10,621 DBS samples were obtained and stored at -20°C. The sample size for our study was calculated based on the previously reported prevalence of HBsAg (10.6%) [11], a precision of 3% with a 95% confidence interval. From the 10,621 DBS, due to insufficient amounts of some samples, only 9,378 were exploitable, covering all 14 prefectures and Bangui, CAR’s capital city. Samples from two prefectures (Nana-Gribizi and Ouham-Pende) were lost during the storage of DBS. Of the 9,378 exploitable DBS, 413 tested HIV-positive, with 8,965 HIV-negative. Among the 8,965 HIV-negative samples, a random selection was performed to include 4,467 samples in this study, to which we added 413 HIV-positive samples. We used all HIV positive samples to determine HIV/hepatitis co-infection prevalence.

### Socio-demographic data

Socio-demographic data were obtained from the Central African Institute of Statistics, Economic and Social Studies (ICASEES), which collected and archived all data related to each sample, including sex, age, ethnicity, educational level, marital status, residential area (region and urban or rural setting). All data were anonymized before processing the analyses. These data were collected from the standard questionnaire of the survey (www.childinfo.org), adapted to the context of Central African Republic.

### Serological tests

Serological tests for hepatitis B, D and C viruses were carried out using the ELISA (enzyme-linked immunosorbent assay) technique on Dried Blood Spot (DBS) stored at the Biological Resources Centre at the Institut Pasteur de Bangui. These samples were made available to the team as soon as the ethical approval was obtained in February 2012. The serological tests began on 8 May 2013. The HBV markers HBsAg, anti-HBc (total), and anti-HBc (IgM) were tested using Abbott-Murex version 3 (Abbott-Murex Biotech Ltd, Dartford, Kent, UK) for the respective markers. Anti-HDV antibody was tested using the DiaSorin kit for HDV, and Anti-HCV antibody (HCV-Ab) using the Murex anti-HCV kit (DiaSorin, DiaSorin S. P.A. UK Branch Central Road). Serological tests were performed and interpreted according to the manufacturer’s recommendations.

### Nucleic Acid Extraction and PCR amplification

Viral nucleic acids (DNA/RNA) were extracted with the Qiagen Kits (QIAamp DNA Blood Mini Kit 250 and QIAamp Viral RNA Mini Kit 50) according to the manufacturer’s procedures. Subsequently, the *PreS1* region of the HBV was amplified using a primer pair P1 (5’-TCACCATATTCTTGGGAACAAGA-3’) /P2 (5’-TTCCTGAACTGGGAGCCACCA-3’) with an expected PCR product of 479 bp. For HIV, a nested RT-PCR was used to amplify the protease region of the *pol* gene using two pairs of primers: PRIn1 (5’-TTT-TTT-AGG-GAA-AAT-TTG-3’) /PRIn2 (5’-ATT-TTC-AGG-CCC-AAT-TTT-TGT-3’) and PRIn3 (5’-CAG-ACC-AGA-GCC-AAC-AGC-3’)/PRIn4 (5’-TCT-TCT-GTC-AAT-GGC-CAT-TGT-3) for primary PCR and nested PCR respectively according to the ANRS protocol (http://www.hivfrenchresistance.org). The R0 region of HDV was amplified with the primers Casey sens (5’CATGCCGACCCGAAGAGGAAAG3’) and anti-sens (5’GAAGGAAGGCCCTCGAGAACAAGA3’), according to the previously published method [16].

### Sequencing

The PCR products were first purified using the Qiagen kit (QIAquick PCR Purification kit, www.qiagen.com/handbooks.), then sent to GATC (Germany) for sequencing using the Sanger method.

### Sequences analysis

The chromatograms of the sequences obtained were cleaned and analyzed in CLC genomic workbench V.8. The HIV subtypes were determined online, using the Stanford University website (HIV DRUG RESISTANCE DATABASE, http://www.sierra2.stanford.edu). For HBV and HCV, the sequences were aligned and compared with the reference sequences downloaded from the database (Genbank), and construction of the phylogenetic trees was carried out using MEGA X software.

All sequences obtained were submitted to NCBI and were given the following accession number: ***HBV*** *(MT155997-MT156072), **HDV** (MT028414-MT028419), and **HIV (**MT121102-MT121107)*.

### Statistical analysis

Data were analyzed using STATA Software.14. Percentages with a 95% confidence interval (CI) were calculated. HBV-positive and HBV-negative groups were compared using χ2 test or Fisher’s exact test for dichotomous variables and Student’s t-test or Wilcoxon rank-sum test for continuous variables.

All variables associated with HBV in univariate analysis (p < 0.25) were then included in a backward stepwise logistic regression. A *p-value* ≤ 0.05 was considered statistically significant.

### Ethical considerations

Because this study was based partly on retrospective data linked to the fourth Multiple Indicator Cluster Survey (MICS-4) carried out in 2010, which was coupled with HIV screening in the general population, the introduction of a new consent form for participants was neither necessary, recommended or indeed possible.

The protocol anonymised the socio-demographic data stored at the « Institut Centrafricain des Statistiques, des Études Économiques et Sociales (ICASEES) » and participants’ blood samples, dried blood spot (DBS), stored at the Biological Resource Centre of the Institut Pasteur de Bangui. The detection of HIV infection during this survey is based on the anonymous-linked protocol developed by the international DHS (Demographic and Health Surveys) programme and approved by the ICF Macro Ethics Committee. According to this protocol, no name or other individual or geographical characteristic that could identify an individual was linked to a blood sample. The standard MICS-4 questionnaire used (www.childinfo.org) was adapted to the context of the Central African Republic. The “Comité Scientifique chargé de Validation des Protocoles et des Résultats de Recherche en Santé (CSVPRS)” of the Faculty of Health Sciences of the University of Bangui (Central African Republic), which acts as the Central African Republic’s national ethics committee, also approved the specific anonymous-linked protocol for the MICS-4 survey under number 2/UB/FACSS/CSVPRS/10. The tests used to detect HIV infection were strictly anonymous, and it was not possible to inform those tested of the results of their examination. However, at the time of the MICS-4, whether or not they had agreed to be tested for HIV, eligible persons (those providing their consent to be included in the survey) received a coupon to obtain, if they so wished, advice and a free HIV test at a “Centre de Prévention et de Dépistage Volontaire (CPDV)”.

The protocol for this study was also assessed by the CSVPRS, which issued a favorable ethical approval under number 6/UB/FACSS/CSVPRS/12. In the protocol submitted to the CSVPRS, we noted the impossibility of collecting informed consent from participants due to the retrospective nature of the study, which used DBS samples stored at the Biological Resources Centre of the Institut Pasteur de Bangui, and the sociodemographic data being anonymous at the outset. We explained that there was no possibility of contacting participants in order to obtain their consent again. The socio-demographic data used to analyse the results of the study were obtained from the ICASEES, which collected and archived all these data in relation to each DBS sample. An anonymous number was used to link the biological results to the individual data, but conserved anonymity, again making it impossible to contact participants to obtain their consent.

## Results

### Population characteristics

Among the 4,317 samples analyzed, 2,297 were from males (53.2%) and 2020 from females (46.8%) with a sex ratio of 1.13. The majority of subjects had either primary school or no education (44.8% and 33.4% respectively). More than half of the participants (2356/4317 or 54.6%) identified themselves as of the Protestant religion. The economic quintiles were second (25.0%), poor (22.7%) and medium (22.7%).

### Prevalence of HBV, HCV, HBV/HIV, and HBV/HDV co-infections

Positive HBsAg and HBc-Ab were found in 555 subjects (12.9%), and 849 (19.7%), respectively. The overall prevalence of anti-HCV was 0.65%. Co-infection of HBV/HIV was found in 49 subjects (1.4%) of 3477 HBV tested population, and HBV/HDV co-infection was found in 92 subjects (16.6%) of the HBV infected population. The prevalence of different hepatitis viral markers and HBV/HIV and HBV/HDV associated with socio-demographic data are reported in *Table 1*.

**Table 1.**
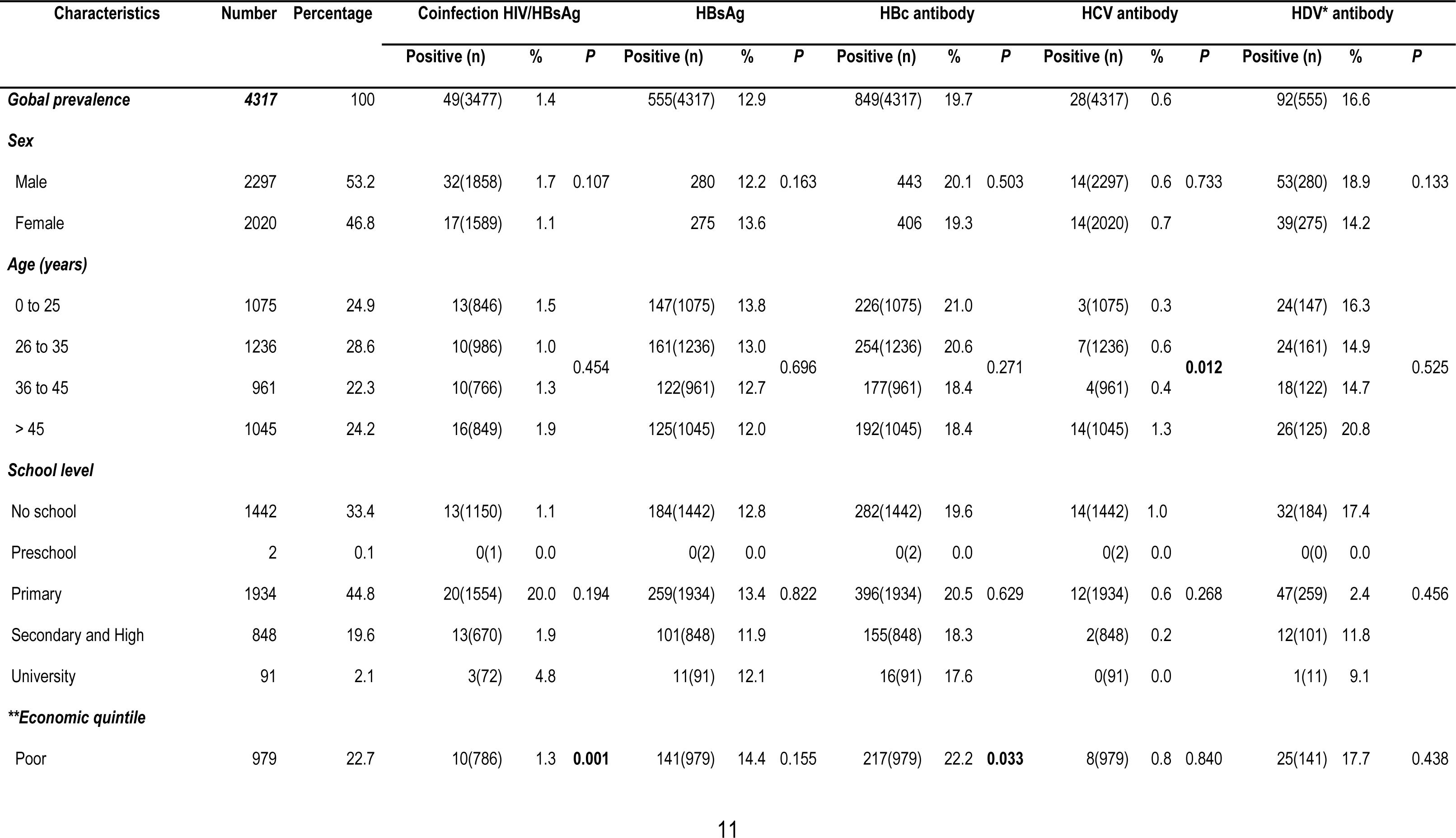
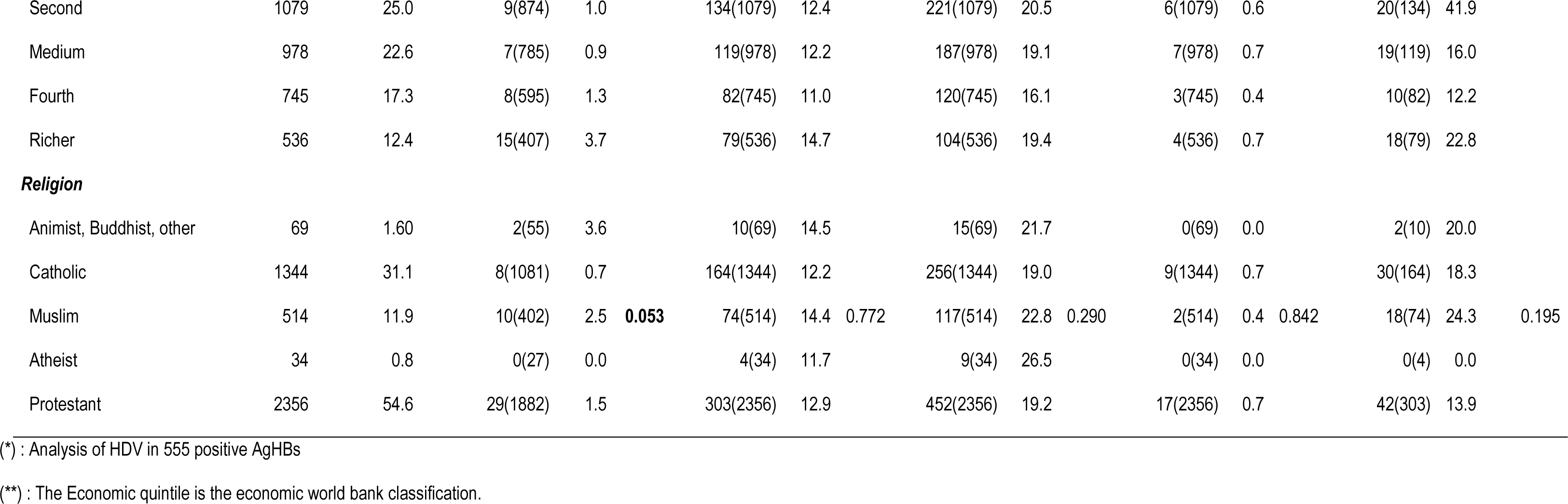
Prevalence and distribution of HBsAg, HBc Ab, HCV, HBsAg/HIV and, HBsAg/HDV coinfections.

No difference of HBV infection was observed between age groups (p=0.70), but for HCV infection, a significant difference was observed for subjects older than 45 years (p=0.012). Among districts the prevalence of the four viruses varied (*Table 2*). Even among districts with the lowest blood samples collection,, Vakaga reported the highest prevalence of HBsAg (33.3%), followed by Bamingui-Bangoran (19.5%) and Ouaka (16.9%). However, in Lobaye and Mambere-Kadéi, HBV prevalence was 8.9% and 8.1% respectively. The global prevalence of HBV/HIV co-infection was 1.41%, with the highest prevalence among Bangui samples (5.1%), followed by Haut-Mbomou (3.7%) and Haute-Kotto (3.4%).

**Table 2.**
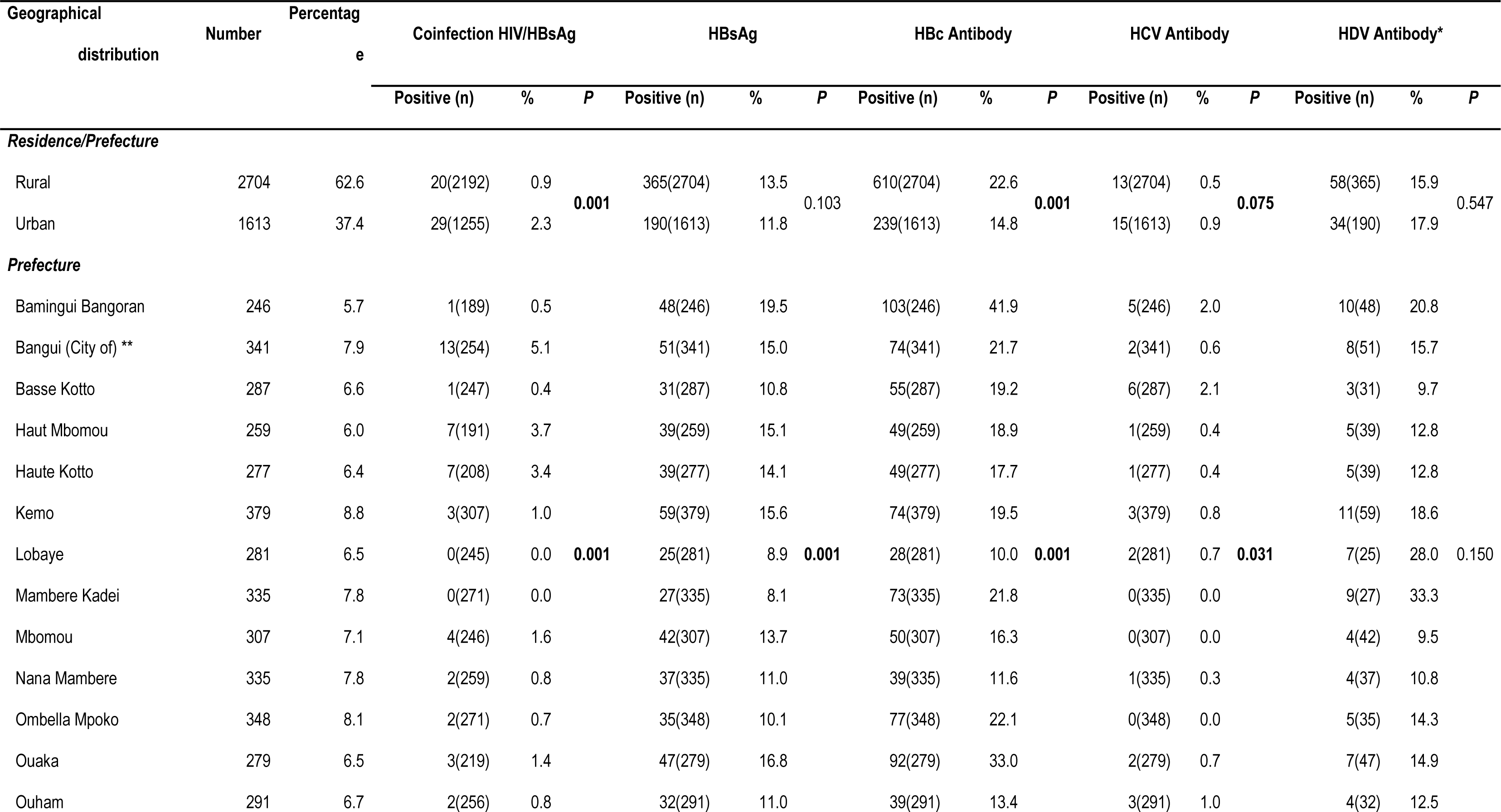
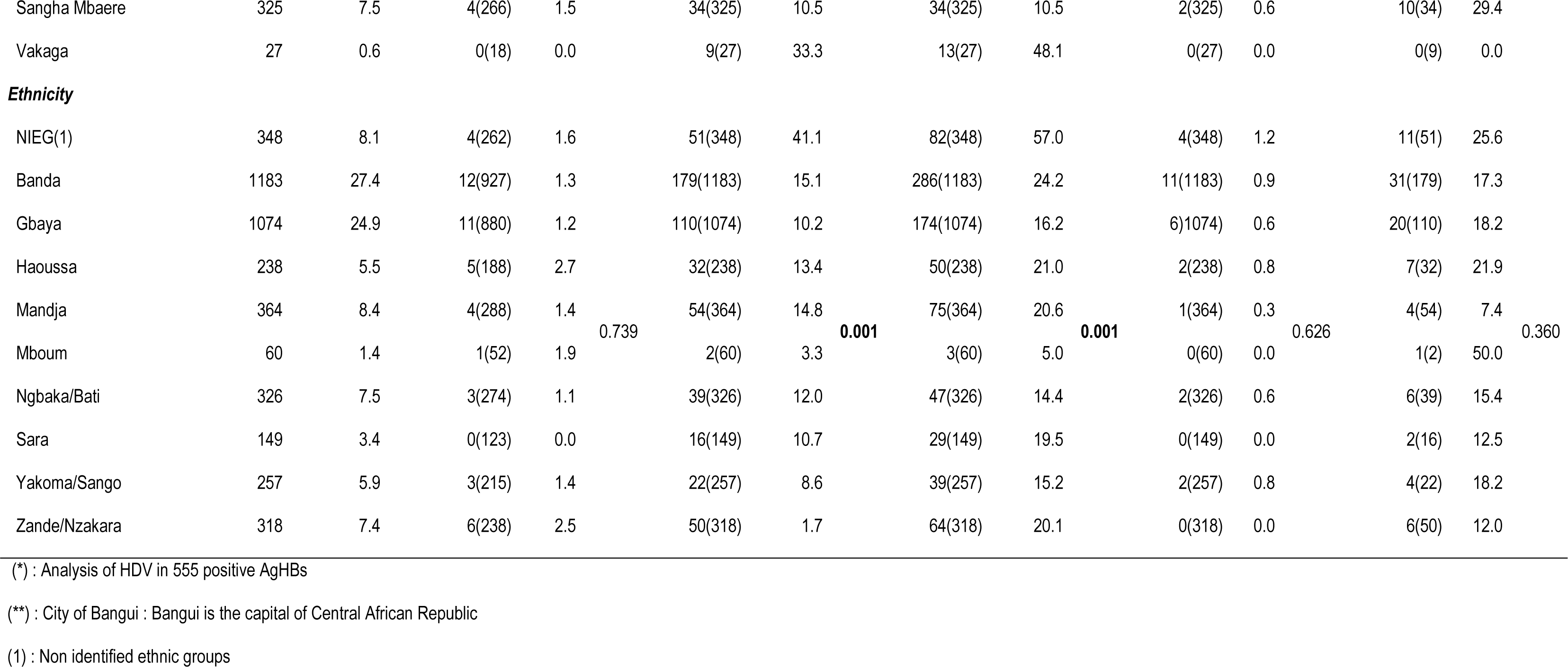
Geographical distribution of hepatitis (B, C and D) viruses.

For ethnic groups, prevalence of HBsAg (41.7%; p< 0.001) and of anti-HBc antibody (57.1%; p< 0.001) was higher among “non identified ethnic groups”, followed by the Banda ethnic group (15.1% and 24.2%), respectively). In contrast, prevalence of HBV/HIV co-infection was slightly higher in the Hausa ethnic group, followed by the Zande/Nzakara ethnic group. Analyses of factors associated with each infection are shown *in Table 3*.

**Table 3.** Association between HBV/HIV coinfection and risk factors.

| Caractéristiques | Co-infection HIV/HBV |  |  |
| --- | --- | --- | --- |
|  | OR | *p* | IC(95%) |
| **\*\*Economic quintile** |  |  |  |
| Wealthier | 4.25 | 0.002 | [1.71 - 10.51] |
| **Residence/Prefecture** |  |  |  |
| Urban | 2.56 | 0.001 | [1.44 - 4.56] |
| **Prefecture** |  |  |  |
| Bangui (City of) ** | 10.14 | 0.026 | [1.31 - 18.21] |
| Vakaga |  |  |  |

### Associated risk factors for HIV-HBV co-infection

In our multivariate analysis, being wealthier (OR 4.25, 95% CI: 1.71-10.51) and living in an urban area (OR 2.56, 95% CI: 1.44 - 4.56), more specifically Bangui and Vakaga (OR 10.14, 95%CI 1.31-78.21), were associated with HIV/HBV co-infection (Table 3).

### Molecular characterization of HBV, HDV, and HIV isolates

A total of 77 HBV strains based on the *PreS1* region were sequenced. Of these 77 strains, 6 were HBV/HDV co-infected and 6 strains were HBV/HIV co-infected. Of the 77 HBV isolates, 72 strains (93.5%) belonged to genotype E, and 5 isolates (6.5%) to genotype D (Figure 2). Of the 92 HDV positive samples, 6 samples were successfully sequenced and all belonged to clade 1 (Figure 3).

**Figure 1:**
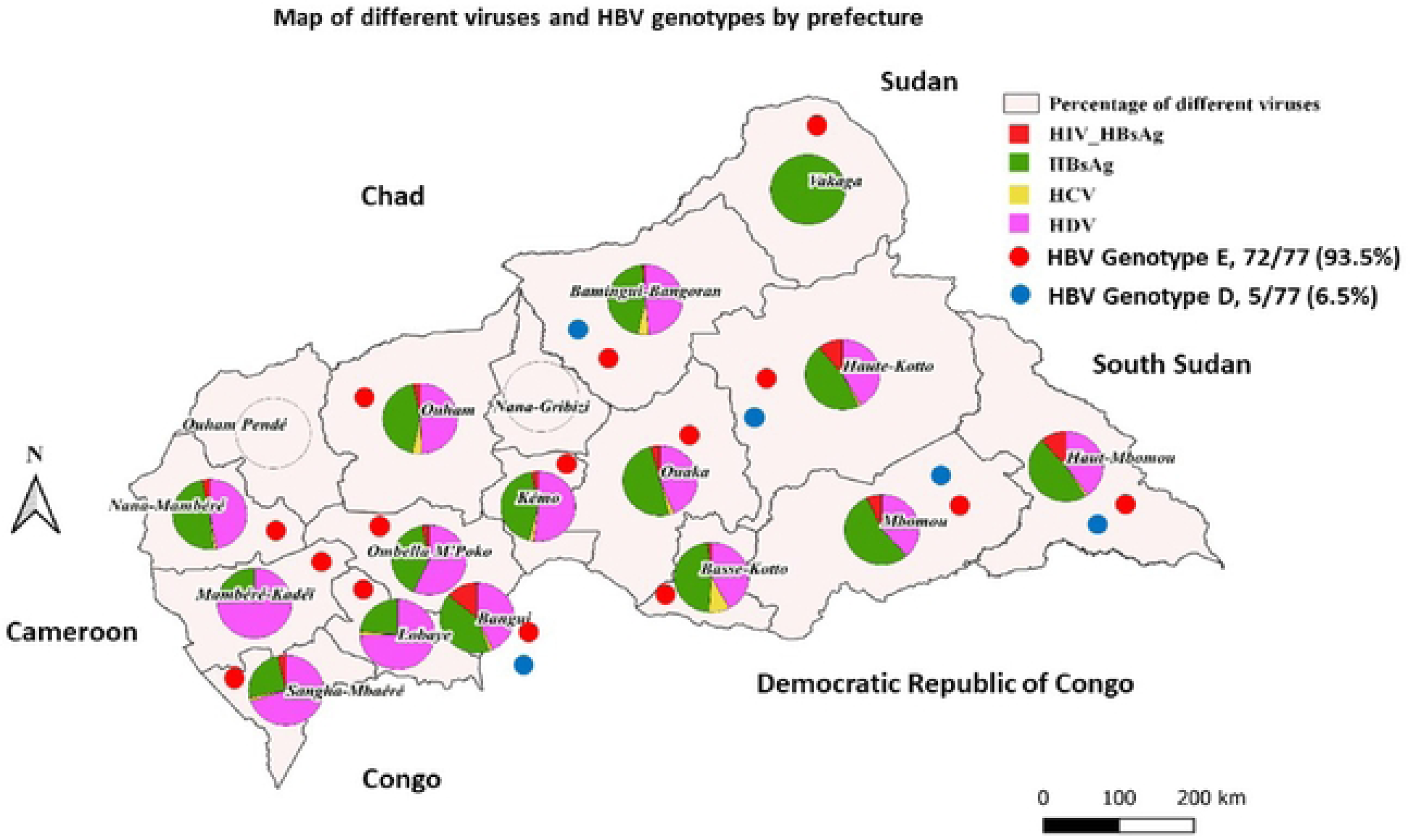
Circulation of viruses and their genotypes in CAR. All four viruses were present in the southeastern part of the Country. Bangui, the capital city, is affected by all three viruses and the circulation of the two genotypes (D and E) of HBV.

**Figure 2:**
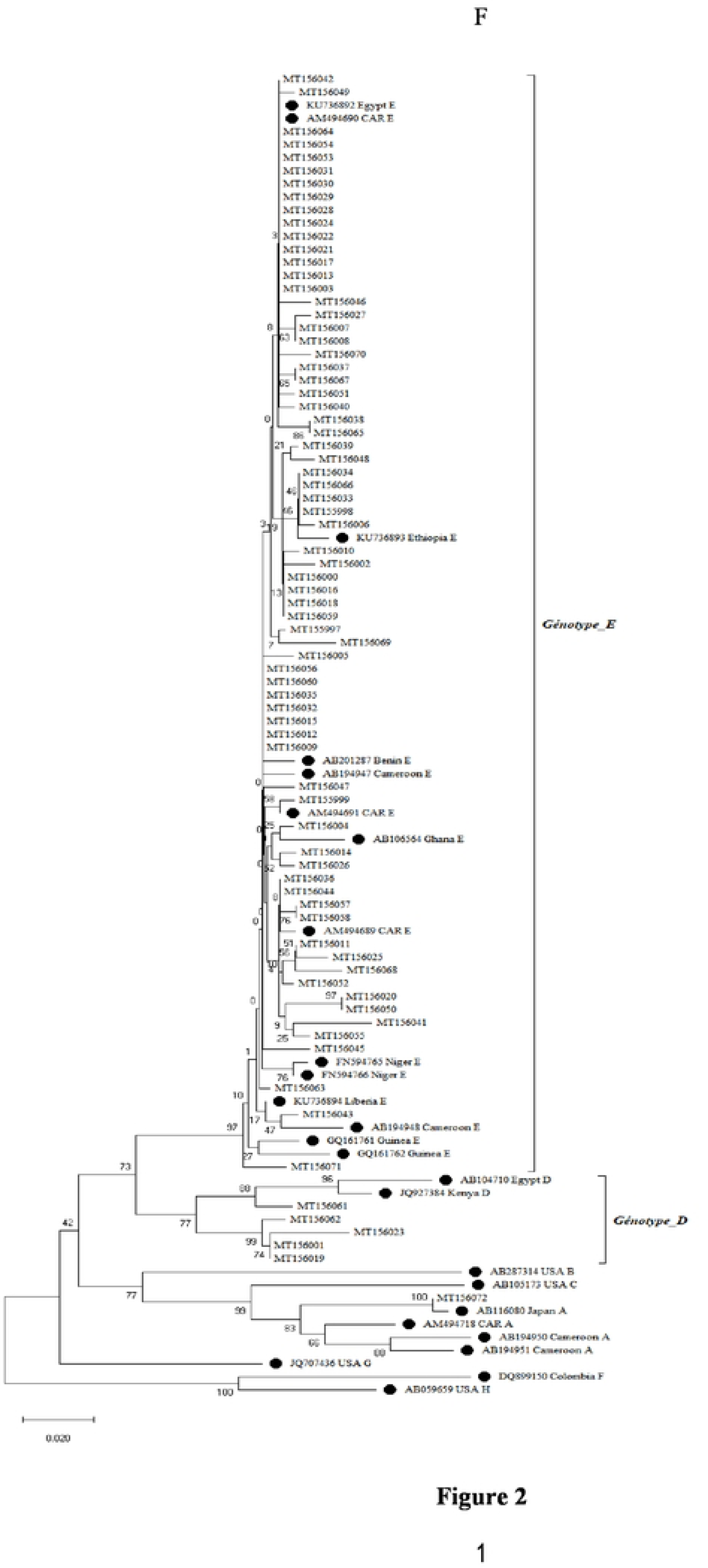
Phylogenetic analyses of 77 HBV isolates. The tree was constructed in MEGA using the Neighbor-joining statistical method, the Kimura-2 parameter model, and the bootstrap method of 1000 replicates. The black cercles represent reference sequence genotypes A-H downloaded from the database. The remaining are HBV sequences discussed in the present study, and the two genotypes (E and D) are mentioned on the tree.

**Figure 3:**
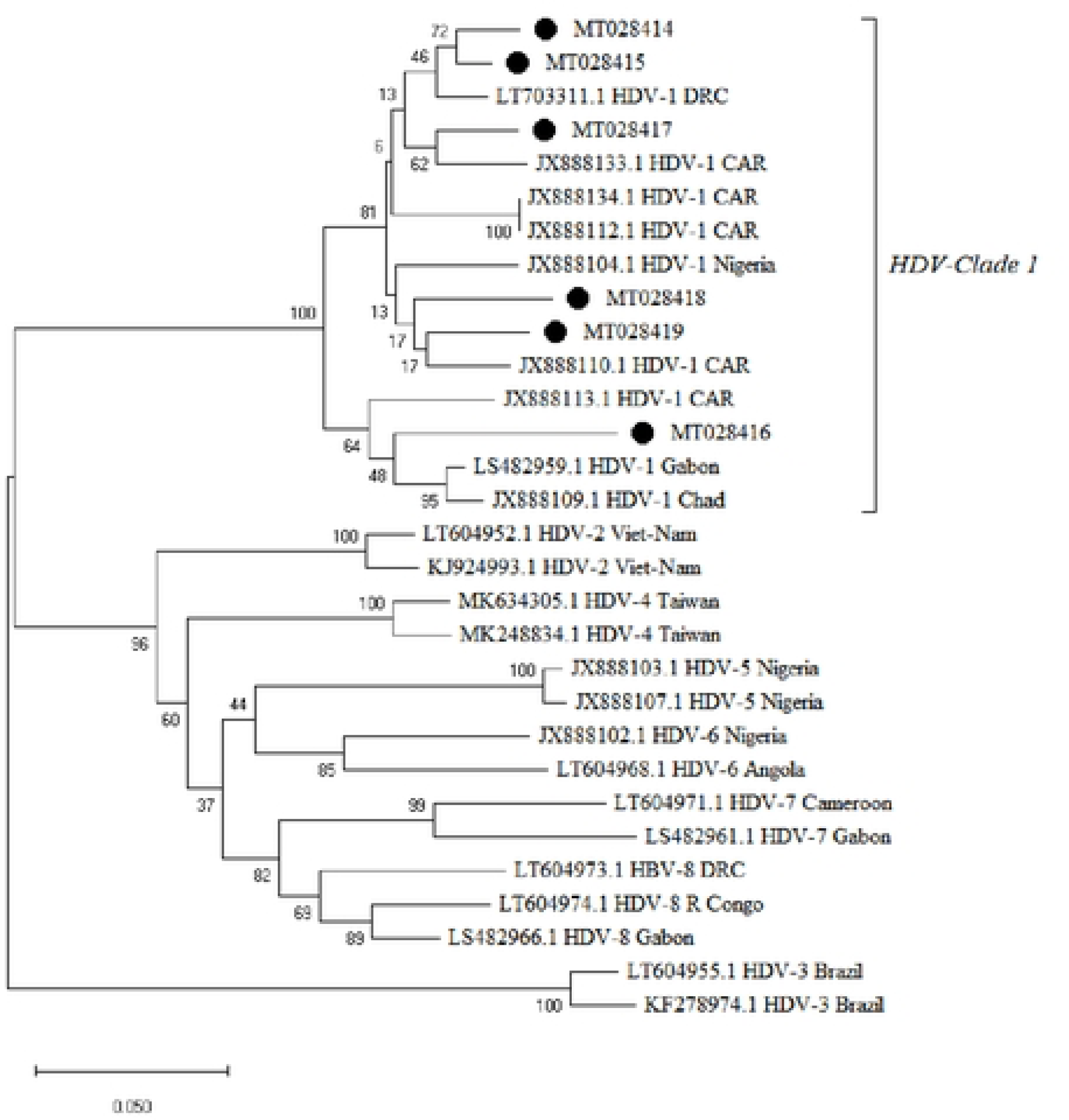
Phylogenetic analyses of the 6 HDV isolates. The tree was constructed in MEGA using the Neighbor-joining statistical method, the Kimura-2 parameter model, and the bootstrap method of 1000 replicates. The black cercles represent the HDV sequences discus by the present study, and the remaining are the reference sequences with the country’s names.

Table 4 presents the 6 sequences of HIV and their GenBank accession number. After similarity analysis via the NCBI database and the Stanford University website sub-types-1 (A1 and G), 4 recombinants of sub-type-1 were identified.

**Table 4.** Different subtypes of HIV-1 isolate.

| *Laboratory ID/ GenBank Accession Number* | *Sub-types* | *Similarity (%)* |
| --- | --- | --- |
| *CARVIHBG280/ MT121104* | CRF01_AE | 100 |
| *CARVIHVK022/ MT121105* | A1 | 100 |
| *CARVIHSM173/ MT121107* | G | 94.6 |
| *CARVIHSM651/ MT121102* | CRF02_AG | 100 |
| *CARVIHKE584/ MT121103* | CRF12_BF | 100 |
| *CAVIHHK175/ MT121106* | CRF11_cpx | 100 |

## Discussion

The study reported the prevalence of viral hepatitis B, C and Delta biomarkers and the molecular characterization of HBV, HDV, HCV and HIV/Hepatitis coinfection of DBS from a nationwide investigation in the CAR.

In all our analyzed cohorts, HBsAg was highly endemic, but its prevalence varied widely among different regions of the CAR, ranging from 8.1% in Mambere Kadéi to 33.3% in Vakaga. Although it is difficult to compare different age groups, HBsAg prevalence was similar in children and adults: 0 to 25 years (13.7%), 25 to 35 years (13.0%), 36 to 45 years (12.7%), and > 45 years (12.0%). These results resemble those of a previous study showing constancy in HBV prevalence among all age groups in the CAR [17]. This finding may indicate a high burden and an excessive risk of early infection with HBV in children, resulting in a high HDV prevalence (16.6%), which has a similar distribution for HBV across all age groups. Previous studies have designated the CAR as an endemic region for HBV and HDV co-infection [11,15,18,19].

The seroprevalence of HBV and HIV co-infection was 1.4%, demonstrating that the CAR is an endemic region for this co-infection.

In this study, the prevalence of HBc antibody was 19.7%. The distribution of this prevalence follows HBsAg prevalence. The presence of these antibodies indicates that a high proportion of the population had been exposed to HBV. That prevalence is high in rural areas (22.6%; p=0.001), among the economically poor (22.7%), in the Vakaga and Bamingui Bangoran regions (48.11 % and 41.9% respectively; p<0.001), and among other ethnic groups as well as the Banda group (57.1 and 24.2%; p<0.001). Compared to previous studies in CAR, this prevalence has significantly decreased, halving since 2010 when it was 42.3% [31], declining to 27.1% in rural communities in 2013 [11], and to 26.7% among students and 21.4% among pregnant women in 2018 [15]. This reduction may be explained by the intensification of hepatitis B surveillance measures in blood transfusion and the introduction of hepatitis B vaccination in the Expanded Program of Immunisation in CAR in 2008.

Although HCV treatment has not been yet initiated in CAR, HCV prevalence in this study was very low (0.65%). Our finding contrasts with a previous review estimating HCV prevalence in Sub-Saharan Africa between 2000 and 2013, which showed West and Central Africa with the highest HCV prevalence [21]. In our study, HCV prevalence in the Bangui samples was 0.59%. We found a higher prevalence in Bamingui-Bangoran samples (2.03%), with variability across regions and no cases at all among Vakaga samples (Table 2). The Bamingui-Bangoran and Vakaga prefectures are in close proximity, but Vakaga is the most sparsely populated prefecture in the CAR, where very few blood samples were collected compared to other prefectures. We would suggest, then, that socio-demographic and economic factors of these prefectoral populations may account for the differential prevalence of these neighboring regions affecting the prevalence of these viruses.

In addition, the prevalence of anti-HCV was lower in the 0 to 25 age group and higher in those over 45 years old. Similar results have been reported in CAR, Cameroon and Gabon, where massive iatrogenic transmission during medical campaigns through improperly sterilized syringes, needles and the transfusion of contaminated blood products may explain the high prevalence among elderly subjects rather than continuous exposure [32, 33].

The main risk factors of HBV identified in our study were economic quintile, religion, residence, prefecture, and ethnicity.

From this study, 77 isolates of HBV were sequenced and genotyped. HBV was categorized into eight genotypes from A to H, based on a divergence of more than 8% in the entire nucleotide sequence of the viral DNA. Two additional genotypes I and J were recently proposed [22]. In the CAR, however, evidence of only three of these HBV genotypes (A, D and E) were reported in previous studies [15,18,23,24].

The present study identified two HBV genotypes (D and E), with HBV genotype E accounting for the dominant strains circulating throughout the CAR (figure 1). This finding conforms with previous studies identifying HBV genotypes E as the most dominant strains circulating in West and Central Africa [18,25]. This HBV genotype has very low diversity and emerged recently, just 200 years or less [25]. HBV genotype D has already been previously described in a Bangui patient with chronic hepatitis [18]. This genotype is predominant in the Maghreb [26], and its low representation in CAR suggests that it may be imported by population and trade movements with other countries in North, West, and Central Africa.

HDV clades (1 to 8) varied across geographic regions [27]. From the present study, all 6 isolates belong to clade 1, in accordance with a previous study [28]. The HDV clade 1 has been reported to have rapid and aggressive HDV virion formation and dissemination, making patients infected with this type suffering more adverse outcomes and decreased chances of survival. This finding requires further study of liver disease patients in the CAR.

Among the 6 isolates of HIV co-infected with HBV in this study, circulating recombinant forms CRF (CRF01_AE, CRF02_AG, CRF12_BF) and the CRF11_cpx complex were identified. These isolates have already been described as part of the pandemic subtypes in Africa [28,29]. In the CAR, the CRF11_cpx complex has been described as predominant. It is associated with antiretroviral resistance, followed by the A1 type [30].

Our study has certain limitations that should be considered, notably the absence of co-infection HIV/HCV, triple infection HB/HCV/HDV or HIV/HBV/HCV, and multiple infection HIV/HBV/HCV/HDV, as well as the absence of detection of HCV-RNA sequences. Additionally, the study did not include an assessment of liver function.

## Conclusion

This study provides a national prevalence and geographical distribution of hepatitis viruses in the CAR, and it shows that hepatitis viral infections remain a serious public health problem in the country. HBV and HDV remain highly prevalent. HBV prevalence was distributed equally across age groups, and genotype E remains dominant. Our results assume? Suggest? that high prevalence of HBV and HBV/HDV co-infection continue to be responsible for morbidity and mortality, despite the introduction of HBV vaccination in 2008. The data and analysis from this study will be useful in assisting authorities to implement measures to limit HBV, HDV, HCV, and HIV infections and coinfection in the CAR.

## Data Availability

All data produced in the present work are contained in the manuscript

## Authors’ contributions

**Conceptualization:** Narcisse Patrice Joseph Komas, Muriel Vray, Alexandre Manirakiza, Sandrine Moussa, Claudine Bekondi

**Data curation:** Narcisse Patrice Joseph Komas, Parvine Basimane-Bisimwa, Brice Martial Yambiyo, Alexandre Manirakiza

**Formal analysis:** Parvine Basimane-Bisimwa, Giscard Koyaweda, Brice Martial Yambiyo, Alexandre Manirakiza, Benjamin Seydou Sombié, Ulrich Vickos, Narcisse Patrice Joseph Komas **Investigation:** Parvine Basimane-Bisimwa, Giscard Koyaweda, Edgarthe Ngaïganam, Ornella Anne Demi Sibiro,

**Methodology:** Parvine Basimane-Bisimwa, Giscard Koyaweda, Muriel Vray, Narcisse Patrice Joseph Komas

**Project Administration :** Narcisse Patrice Joseph Komas,

**Resources:** Sandrine Moussa, Pulchérie Pélembi, Narcisse Patrice Joseph Komas

**Supervision:** Narcisse Patrice Joseph Komas

**Writing – original draft:** Parvine Basimane-Bisimwa, Giscard Koyaweda, Narcisse Patrice Jospeh Komas

**Writing – review & editing:** Parvine Basimane-Bisimwa, Giscard Wilfried Koyaweda, Ulrich Vickos, Tamara Giles-Vernick, Alexandre Manirakiza, Muriel Vray, Narcisse Patrice Komas

## References

[1] Valenzuela P. Hepatitis A, B, C, D and E viruses: Structure of their genomes and general properties. Gastroenterol Jpn 1990;25:62–71. doi:10.1007/BF02779931.

[2] Jefferies M, Rauff B, Rashid H, Lam T, Rafiq S. Update on global epidemiology of viral hepatitis and preventive strategies. World J Clin Cases 2018;6:589–99. doi:10.12998/wjcc.v6.i13.589.

[3] Papastergiou V, Lombardi R, MacDonald D, Tsochatzis EA. Global epidemiology of hepatitis B virus (HBV) infection. Curr Hepat Rep 2015;14:171–8. doi:10.1007/s11901-015-0269-3.

[4] WHO. World Health Organization. Global Hepatitis Report. 2017a. Glob Hepat Program Dep HIV/AIDS 2017;Disponível:14.

[5] Ott JJ, Stevens GA, Groeger J, Wiersma ST. Global epidemiology of hepatitis B virus infection: New estimates of age-specific HBsAg seroprevalence and endemicity. Vaccine 2012;30:2212–9. doi:10.1016/j.vaccine.2011.12.116.

6. [6] Hepatitis D n.d. https://www.who.int/news-room/fact-sheets/detail/hepatitis-d (accessed June 19, 2020).

[7] Abbas Z, Qureshi M, Hamid S, Jafri W. Hepatocellular carcinoma in hepatitis D: does it differ from hepatitis B monoinfection? Saudi J Gastroenterol 2012;18:18–22. doi:10.4103/1319-3767.91731.

[8] Puigvehí M, Moctezuma-Velázquez C, Villanueva A, Llovet JM. The oncogenic role of hepatitis delta virus in hepatocellular carcinoma. JHEP Reports 2019;1:120–30. 10.1016/j.jhepr.2019.05.001.

[9] Abbas Z, Abbas M, Abbas S, Shazi L. Hepatitis D and hepatocellular carcinoma. World J Hepatol 2015;7:777–86. doi:10.4254/wjh.v7.i5.777.

[10] Hoffmann CJ TC. Clinical implications of HIV and hepatitis B co-infection in Asia and Africa. Lancet Infect Dis 2007:402–409.jour

[11] Stockdale AJ, Chaponda M, Beloukas A, Phillips RO, Matthews PC, Papadimitropoulos A, et al. Prevalence of hepatitis D virus infection in sub-Saharan Africa: a systematic review and meta-analysis. Lancet Glob Heal 2017;5:e992–1003. doi:10.1016/S2214-109X(17)30298-X.

[12] Komas NP, Vickos U, Hübschen JM, Béré A, Manirakiza A, Muller CP, et al. Cross-sectional study of hepatitis B virus infection in rural communities, Central African Republic. BMC Infect Dis 2013;13:286. doi:10.1186/1471-2334-13-286.

[13] Crovari P, Santolini M, Bandettini R, Bonanni P, Branca P, Coppola RC. Epidemiology of HBV and HDV infections in a rural area of Central African Republic. Prog Clin Biol Res 1991;364:69–73.

14. [14] Classement des États du monde par prévalence du VIH (% population âgée de 15 à 49 ans) n.d. https://atlasocio.com/classements/sante/maladies/classement-etats-par-prevalence-vih-pourcentage-monde.php#google_vignette (accessed January 18, 2021).

[15] Komas NP, Ghosh S, Abdou-Chekaraou M, Pradat P, Al Hawajri N, Manirakiza A, et al. Hepatitis B and hepatitis D virus infections in the Central African Republic, twenty-five years after a fulminant hepatitis outbreak, indicate continuing spread in asymptomatic young adults. PLoS Negl Trop Dis 2018;12:e0006377. doi:10.1371/journal.pntd.0006377.

[16] Ivaniushina V, Radjef N, Alexeeva M, Gault E, Semenov S, Salhi M, et al. Hepatitis delta virus genotypes I and II cocirculate in an endemic area of Yakutia, Russia. J Gen Virol 2001;82:2709–18. doi:10.1099/0022-1317-82-11-2709.

[17] Andernach IE, Leiss L V., Tarnagda ZS, Tahita MC, Otegbayo JA, Forbi JC, et al. Characterization of hepatitis delta virus in sub-Saharan Africa. J Clin Microbiol 2014;52:1629–36. doi:10.1128/JCM.02297-13.

[18] Bekondi C, Olinger CM, Boua N, Talarmin A, Muller CP, Le Faou A, et al. Central African Republic is part of the West-African hepatitis B virus genotype E crescent. J Clin Virol 2007;40:31–7. doi:10.1016/j.jcv.2007.05.009.

[19] Bekondi C. Aspects cliniques et epidemiologiques des infections a virus de l’hepatite b en republique centrafricaine. Université Henri Poincaré - Nancy 1, 2007. doi:2008NAN10129.

[20] Bertoletti A, Hong M. Age-Dependent Immune Events during HBV Infection from Birth to Adulthood: An Alternative Interpretation. Front Immunol 2014;5:441. doi:10.3389/fimmu.2014.00441.

[21] Mora N, Adams WH, Kliethermes S, Dugas L, Balasubramanian N, Sandhu J, et al. A Synthesis of Hepatitis C prevalence estimates in Sub-Saharan Africa: 2000-2013. BMC Infect Dis 2016;16:1–8. doi:10.1186/s12879-016-1584-1.

[22] Lin CL KJ. The clinical implications of hepatitis B virus genotype: recent advances. J Gastroenterol Hepatol 2011;26:123–130.

[23] Toxoplasmosis of Animals and Man. By J. P. Dubey and C. P. Beattie. 220 pages. ISBN 0 8493 4618 5. CRC Press, Boca Raton, 1988. £108.00. Parasitology 1990;100:500–1. doi:10.1017/s0031182000078914.

[24] Koyaweda Giscard Wilfried, Rose J, Machuka E, Juma J, Macharia R, Patrice N, et al. Detection of circulating hepatitis B virus immune escape and polymerase mutants among HBV-positive patients attending Institut Pasteur de Bangui, Central African Republic. Int J Infect Dis 2020;90:138–44. doi:10.1016/j.ijid.2019.10.039.

[25] Kramvis A. Genotypes and genetic Variability of Hepatitis B virus. Intervirology 2014;57:141–50. doi:10.1159/000360947.

[26] Forbi JC, Ben-Ayed Y, Xia GL, Vaughan G, Drobeniuc J, Switzer WM KY. Disparate distribution of hepatitis B virus genotypes in four sub-Saharan African countries. J Clin Virol 2013;1:59–66. doi:doi: 10.1016/j.jcv.2013.06.028.

[27] Le Gal F, Gault E, Ripault MP, Serpaggi J, Trinchet JC, Gordien E, et al. Eighth major clade for hepatitis delta virus. Emerg Infect Dis 2006;12:1447–50. doi:10.3201/eid1209.060112.

[28] Brennan CA, Bodelle P, Coffey R, Devare SG, Golden A, Hackett J, et al. The Prevalence of Diverse HIV-1 Strains Was Stable in Cameroonian Blood Donors From 1996 to 2004 2008;49:432–9.

[29] Ndembi N, Takehisa J, Zekeng L, Kobayashi E. Genetic Diversity of HIV Type 1 in Rural Eastern Cameroon 2004;37:1641–50.

[30] Moussa S, Pinson P, Pelembi P, Gody J-C, Mbitikon O, Fikouma V, et al. First data on HIV-1 resistance mutations to antiretroviral drugs in Central African Republic. AIDS Res Hum Retroviruses 2010;26:1247–8. doi:10.1089/aid.2010.0091.

[31] Komas NP, Baï-Sepou S, Manirakiza A, Léal J, Béré A, Le Faou A. The prevalence of hepatitis B virus markers in a cohort of students in Bangui, Central African Republic. BMC Infect Dis. 2010;10:226. Published 2010 Jul 29. doi:10.1186/1471-2334-10-226.

[32] Njouom, R., Frost, E., Deslandes, S., Mamadou-Yaya, F., Labbe, A. C., Pouillot, R., … & Pepin, J. Predominance of hepatitis C virus genotype 4 infection and rapid transmission between 1935 and 1965 in the Central African Republic. Journal of general virology. 2009; 90(10), 2452–2456.

[33] Njouom, R., Pasquier, C., Ayouba, A., Gessain, A., Froment, A., Mfoupouendoun, J., … & Nerrienet, E. High rate of hepatitis C virus infection and predominance of genotype 4 among elderly inhabitants of a remote village of the rain forest of South Cameroon. Journal of medical virology. 2003 ; 71(2), 219–225.

